# Multinational Prevalence of Neurological Phenotypes in Patients Hospitalized with COVID-19

**DOI:** 10.1101/2021.01.27.21249817

**Authors:** Trang T. Le, Alba Gutiérrez-Sacristán, Jiyeon Son, Chuan Hong, Andrew M. South, Brett K. Beaulieu-Jones, Ne Hooi Will Loh, Yuan Luo, Michele Morris, Kee Yuan Ngiam, Lav P. Patel, Malarkodi J. Samayamuthu, Emily Schriver, Amelia LM Tan, Jason Moore, Tianxi Cai, Gilbert S. Omenn, Paul Avillach, Isaac S. Kohane, 4CE Consortium, Shyam Visweswaran, Danielle L. Mowery, Zongqi Xia

**Author notes:** Corresponding Authors: Zongqi Xia, MD, PhD, Department of Neurology, University of Pittsburgh, Biomedical Science Tower 3, Suite 7014, 3501 5^th^ Avenue, Pittsburgh, PA 15260. Drs. Le, Gutiérrez-Sacristán and Son share co-first authorship. Drs. Visweswaran, Mowery and Xia share co-senior authorship. 4CE Consortium: International Consortium for Clinical Characterization of COVID-19 by EHR.

## Abstract

**OBJECTIVE:** Neurological complications can worsen outcomes in COVID-19. We defined the prevalence of a wide range of neurological conditions among patients hospitalized with COVID-19 in geographically diverse multinational populations.

**METHODS:** Using electronic health record (EHR) data from 348 participating hospitals across 6 countries and 3 continents between January and September 2020, we performed a cross-sectional study of hospitalized adult and pediatric patients with a positive SARS-CoV-2 reverse transcription polymerase chain reaction test, both with and without severe COVID-19. We assessed the frequency of each disease category and 3-character International Classification of Disease (ICD) code of neurological diseases by countries, sites, time before and after admission for COVID-19, and COVID-19 severity.

**RESULTS:** Among the 35,177 hospitalized patients with SARS-CoV-2 infection, there was increased prevalence of disorders of consciousness (5.8%, 95% confidence interval [CI]: 3.7%-7.8%, *p*_FDR_<.001) and unspecified disorders of the brain (8.1%, 95%CI: 5.7%-10.5%, *p*_FDR_<.001), compared to pre-admission prevalence. During hospitalization, patients who experienced severe COVID-19 status had 22% (95%CI: 19%-25%) increase in the relative risk (RR) of disorders of consciousness, 24% (95%CI: 13%-35%) increase in other cerebrovascular diseases, 34% (95%CI: 20%-50%) increase in nontraumatic intracranial hemorrhage, 37% (95%CI: 17%-60%) increase in encephalitis and/or myelitis, and 72% (95%CI: 67%-77%) increase in myopathy compared to those who never experienced severe disease.

**INTERPRETATION:** Using an international network and common EHR data elements, we highlight an increase in the prevalence of central and peripheral neurological phenotypes in patients hospitalized with SARS-CoV-2 infection, particularly among those with severe disease.

## Introduction

The World Health Organization declared coronavirus disease 2019 (COVID-19) due to the severe acute respiratory syndrome coronavirus 2 (SARS-CoV-2) infection as a global pandemic on March 11, 2020.^1^ Increasing evidence points to the multi-organ involvement of COVID-19, including the nervous system, which increases morbidity and mortality. Understanding the neurological manifestations is essential for developing strategies to mitigate disability and death from the neurological consequences of COVID-19.

Many case reports and reviews have highlighted neurological phenotypes in adults with COVID-19, including cerebrovascular disease^2^, meningoencephalitis and encephalomyelitis^3,4^, encephalopathy^5^, headache^6^, cranial neuropathies^7^, Guillain-Barré syndrome^8,9^, plexopathy^10^, anosmia and ageusia^11,12^, and cognitive dysfunction^13^. Children with COVID-19 have similar presentations, including ischemic stroke, encephalopathy, headache, and muscle weakness^14,15^. Thus, neurological complications in the setting of COVID-19 affect both the central and the peripheral nervous system. Prior studies reported the prevalence of COVID-19-associated neurological conditions at country level, including China^16^, the United Kingdom^17^, and Italy^18^, but few studies have used validated common data elements to examine the prevalence of neurologic conditions across countries. While studies have described the spectrum of neurological conditions associated with severe COVID-19^19–21^, no multinational study has compared the prevalence of neurological complications in patients with severe versus non-severe disease, based on respiratory and/or critical illness status. Given the health consequences of neurological complications, recognizing the associated neurological complications could inform prevention, diagnosis, and treatment.

Clinical data in electronic health records (EHRs) enable studies across institutions to estimate the prevalence of neurological conditions in patients with COVID-19. There are several challenges when using international and multi-institutional EHR data for clinical discovery: (1) adopting common data elements and standardization processes for representing and aggregating clinical events, (2) sharing clinical data across institutions while adhering to multi-national patient privacy laws such as the United States Health Insurance Portability and Accountability Act (HIPAA) and the European Union General Data Protection Regulation (GDPR), and (3) rapidly developing and deploying analytical solutions at scale by leveraging existing informatics infrastructures and frameworks. Our team created the International Consortium for Clinical Characterization of COVID-19 by EHR (4CE; www.covidclinical.net), standardizing and aggregating EHR data from nearly 350 hospitals across six countries to identify and address critical clinical and epidemiological questions relevant to COVID-19.^22,23^ Leveraging the highly scalable, multinational networks of the 4CE consortium, we estimated the prevalence of neurological conditions in hospitalized patients with reverse transcription polymerase chain reaction (PCR)-confirmed SARS-CoV-2 infection by site and country, and compared the differences in the prevalence of neurological conditions between patients with and those without severe disease based on the internationally validated 4CE criteria.^24^

## Methods

### Patients and Data

4CE contributing sites began in March 2020 to collect EHR data from hospitalized patients with positive SARS-CoV-2 RT-PCR tests. The analyzed data spanned from January 2020 through early September 2020. We defined COVID-19-related hospitalization as the first hospital admission that occurred between 7 days before and up to 14 days after the first positive SARS-CoV-2 PCR test. The first admission date within this −7 to +14 day window is the index admission date.

According to the 4CE consortium agreement, we de-identified contributing sites to protect site confidentiality. The institutional review board of each participating site approved the sharing of anonymous, aggregate data in compliance with multi-national patient privacy laws exempting the requirement for individual patient consent. A small level of obfuscation to preserve site-specific privacy and to reduce the risk of patient re-identification may slightly but not significantly affect the total patient counts.

Using the standardized 4CE common data model^22,23^, we collected demographics (age, sex, self-identified race/ethnicity) and the International Classification of Disease (ICD) codes (versions 9 or 10) pertaining to neurological conditions (**Fig. 1**) as well as COVID-19 severity, defined according to the 4CE definition.^24^ Of the 45 contributing sites, five sites (all from Italy) provided only ICD-9 codes while the remaining 40 sites provided predominantly ICD-10 codes. As such, we used ICD-10 data for the main analyses and ICD-9 data for supplementary analyses. We used the first three alphanumeric characters of a given ICD code, which designates the category of the disease or injury (*e.g.,* ICD-10 G44 denotes the category of “Other headache syndromes”).

**Figure 1.**
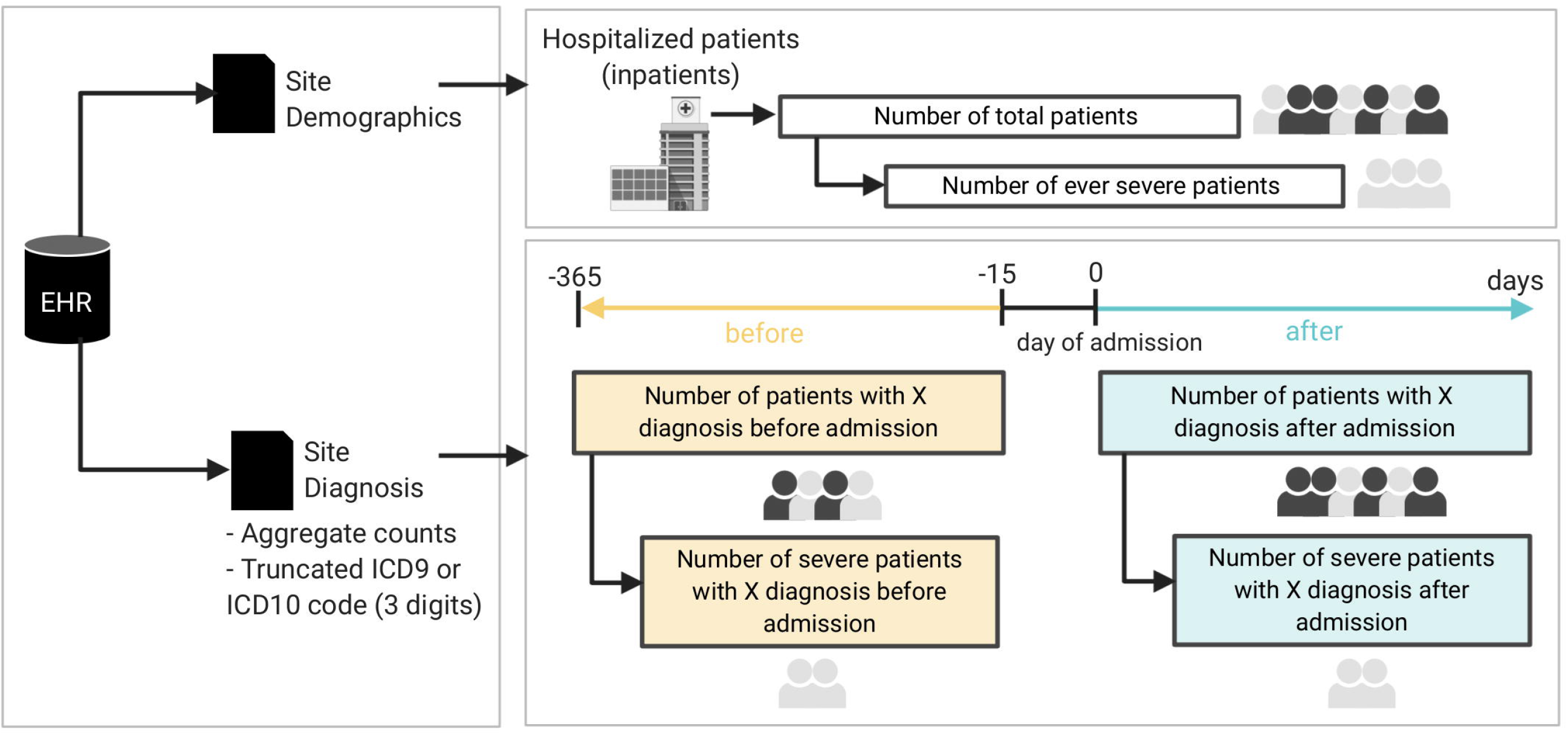
Schematic diagram of the cohort and data generation workflow for each site.

For all patients hospitalized with COVID-19, we collected ICD codes at two time periods, *before* and *after* the admission date. The period before COVID-19 hospitalization ranged from - 365 days to −15 days preceding the admission date. Inclusion of EHR data up to one year before admission is a pragmatic decision to balance available data from all sites and minimize past medical conditions that might not be relevant. The period after admission date ranged from the date of admission to the end of the hospitalization. According to the pre-planned consortium-wide strategy, we excluded all codes in the two weeks preceding the hospital admission date to ensure that diagnoses before admission were independent of COVID-19. It could take a few days from SARS-CoV-2 infection to symptom onset and additional days before a positive PCR test and/or hospital admission. Similarly, we analyzed ICD codes before and after the admission date according to whether patients ever met the 4CE criteria for severe COVID-19^24^ (**Fig. 1**).

### Exposures

We first examined all hospitalized patients with positive SARS-CoV-2 PCR tests. We then examined hospitalized patients with COVID-19 who met any of the definitions for severe COVID-19^24^, including advanced respiratory care management at any point during their hospitalization. Definitions of severe COVID-19 included diagnoses, procedures, laboratory results, or medications (**Table 1**).

**Table 1.** Definition of severe COVID-19 based on the 4CE Severity Criteria.

| Severe Illness Category | Clinical Events |
| --- | --- |
| Diagnoses | Acute respiratory distress syndrome, ventilator-associated pneumonia |
| Procedures | Insertion of endotracheal tube; invasive mechanical ventilation |
| Laboratory results | PaCO<sub>2</sub>, PaO<sub>2</sub> |
| Medications | General anesthetics; benzodiazepine derivatives; muscle relaxants; other hypnotics and sedatives; adrenergic and dopaminergic agents; other cardiac stimulants; other respiratory system products; phosphodiesterase inhibitors; platelet aggregation inhibitors excluding heparin; vasopressin and analogues |

Please see detail of the 4CE severity criteria in a separate manuscript (accepted at Journal of American Medical Informatics):
https://www.medrxiv.org/content/10.1101/2020.10.13.20201855v1.full

### Neurological Outcomes

We queried 21 ICD-10 codes in 12 categories pertaining to neurological phenotypes following a literature search in August 2020. The neurological disease categories included consciousness, coordination, dizziness, headache, inflammatory, muscle, neuropathy, psychiatric, seizure, vascular, vision and other unspecified neurological conditions (**Table 2**).

**Table 2.** Mapping of neurological disease categories to ICD-10 category codes and their descriptions.

| **Disease Category** | **ICD-10 Code** | **ICD-10 Code Description** |
| --- | --- | --- |
| Consciousness | R41 | Other symptoms and signs involving cognitive functions and awareness |
| Coordination | R27 | Other lack of coordination |
| Dizziness | R42 | Dizziness and giddiness |
| Headache | G44 | Other headache syndromes |
| Inflammatory | G03 | Meningitis due to other and unspecified causes |
| Inflammatory | G04 | Encephalitis, myelitis and encephalomyelitis |
| Muscle | G72 | Other and unspecified myopathies |
| Muscle | M60 | Myositis |
| Neuropathy | G61 | Inflammatory polyneuropathy |
| Neuropathy | G65 | Sequelae of inflammatory and toxic polyneuropathies |
| Neuropathy | R43 | Disturbances of smell and taste |
| Other | G93 | Other disorders of the brain |
| Psychiatric | F29 | Unspecified psychosis not due to a substance or known physiological condition |
| Seizure | G40 | Epilepsy and recurrent seizures |
| Vascular | G45 | Transient cerebral ischemic attacks and related syndromes |
| Vascular | G46 | Vascular syndromes of brain in cerebrovascular disease |
| Vascular | I60 | Nontraumatic subarachnoid hemorrhage |
| Vascular | I61 | Nontraumatic intracerebral hemorrhage |
| Vascular | I62 | Other and unspecified nontraumatic intracranial hemorrhage |
| Vascular | I67 | Other cerebrovascular diseases |
| Vision | H54 | Blindness and low vision |

### Statistical Analyses

We first compared the prevalence of each neurological ICD code and disease category among all hospitalized patients with COVID-19. For each ICD code, we reported the total count and the proportion of patients hospitalized with COVID-19 at each site (and each country), both before and after admission date (**Supplementary Material, eEq. 1**). We used the proportion data before admission as reference control. We calculated the difference in the proportion of cases with a given ICD code before and after admission date (**eEq. 2**) and used paired two-sided t-tests to examine whether there was a statistically significant difference in the proportion after admission date when compared to before admission date.

We next compared the prevalence of each neurological ICD code and disease category after admission date between patients who ever met the criteria of severe COVID-19 and those who did not, by site and by country. For each ICD code, we computed the expected number of severe cases (**eEq. 3**) and compared with the observed number of severe cases for the given neurological code. To examine the difference in proportion of severe cases for the neurological ICD codes, we calculated the enrichment of each neurological ICD code by dividing the observed number of severe cases by the expected number of severe cases and reported a value of log_2_ enrichment (LOE) and its 95% confidence interval (CI) (**eEq. 4**). We estimated the LOE 95% CI using the Delta method.^25^ Finally, we computed the *p*-values using Fisher’s exact test^26^ and corrected for multiple hypothesis testing with Benjamini–Hochberg’s false discovery rate (FDR) procedure.^27^ A result was statistically significant if *p*_FDR_<0.05.

### Data / Code Availability

The 4CE consortium does not have permission from each individual contributing site to release EHR data for public access. Our analysis results are available in browsable R notebooks (https://github.com/covidclinical/Phase1.1NeuroRCode/) under CC BY 4.0, with source code distributed under a BSD 3-Clause License.

## Results

### Demographics

Among the 35,177 hospitalized patients with PCR-confirmed SARS-CoV-2 infection, aggregate demographic data were available for 34,647 patients (98.5%) from 348 hospitals (affiliated with 43 sites, after excluding two US sites with incomplete demographic data). The cohort had a greater proportion of men (20,814, 60.1%) than women (13,546, 39.1%), while 287 (0.8%) patients had unknown sex (**Fig. 2**). Variation in the proportions of severe COVID-19 cases among the US sites was large, with 18 sites reporting >90% severe cases. The proportions of severe COVID-19 cases were higher in France than Germany, Italy, Spain, and Singapore. There was no clear relationship between COVID-19 severity and median age (**Fig. 2B**). Most sites in Europe did not report race. Among the US sites, there was a disproportionately high proportion of Black individuals (**Fig. 2C**). The study population included a high proportion of individuals above age 50 years and a low proportion of children (<age 18 years) (**Fig. 2D**).

**Figure 2.**
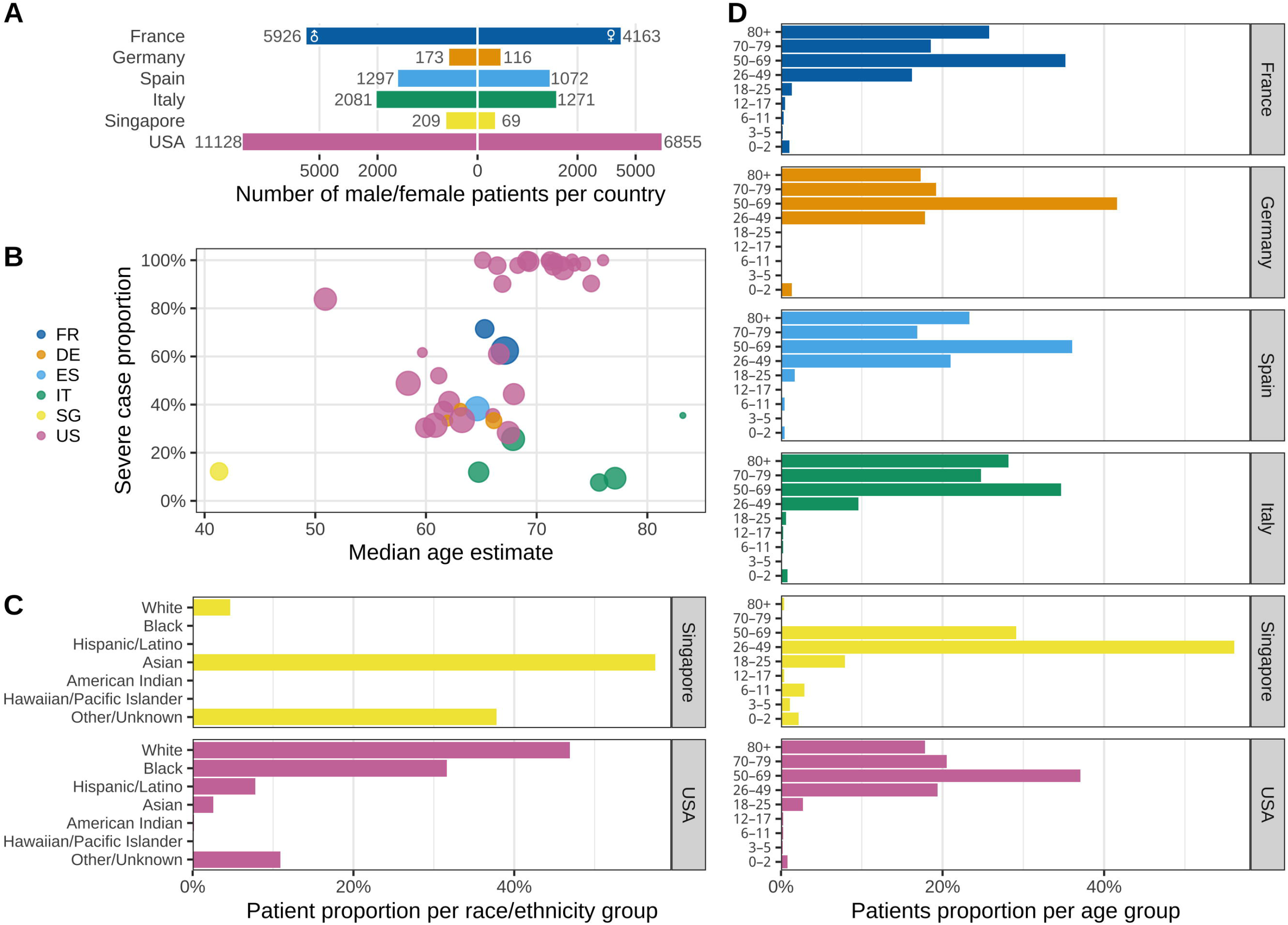
Characteristics of the study population across sites and countries. (**A**) Total number of male (left) and female (right) patients grouped by country shown in square-root scale. (**B**) Proportion of ever-severe cases by median age estimate at each site, grouped by country. Node size corresponds to the total number of patients per site. (**C**) Distribution of self-identified race among patients at sites in Singapore and the United States. The Other/Unknown category includes patients who did not identify with any of the predefined race categories and/or whose data were not reported. Most European sites did not report race. (**D**) Average proportion of patients in each age group within each country. FR, France; DE, Germany; ES, Spain; IT, Italy; SG, Singapore; US(A), United States of America.

### Prevalence of Neurological Conditions in All Patients

Using the prevalence before admission as reference, we found that the majority of contributing sites (77%) reported an increase in the proportion of hospitalized COVID-19 patients with disorders of consciousness (ICD-10 R41: “Other symptoms and signs involving cognitive functions and awareness”) with a mean increase of 5.8% (95% CI: 3.7-7.8%, *p*_FDR_<0.001) after admission (**Fig. 3, eFig. 1**). Similarly, 84% of sites reported an increase in the proportion of patients with “Unspecified disorders of the brain” (ICD-10 G93, including “encephalopathy”) with a mean increase of 8.1% (5.7-10.5%, *p*_FDR_<0.001) after admission. (see <u>interactive data repository</u>:
https://covidclinical.github.io/Phase1.1NeuroRCode/01-analysis-icd10.html#prevalence-change-table). The proportion of patients with “epilepsy and recurrent seizures” (ICD-10 G40), “encephalitis, myelitis, and encephalomyelitis” (ICD-10 G04) and “other and unspecified myopathies” (ICD-10 G72) increased after admission, but these findings were not significant after adjusting for multiple testing. Likewise, none of the other neurological conditions showed a statistically significant difference in prevalence after admission.

**Figure 3.**
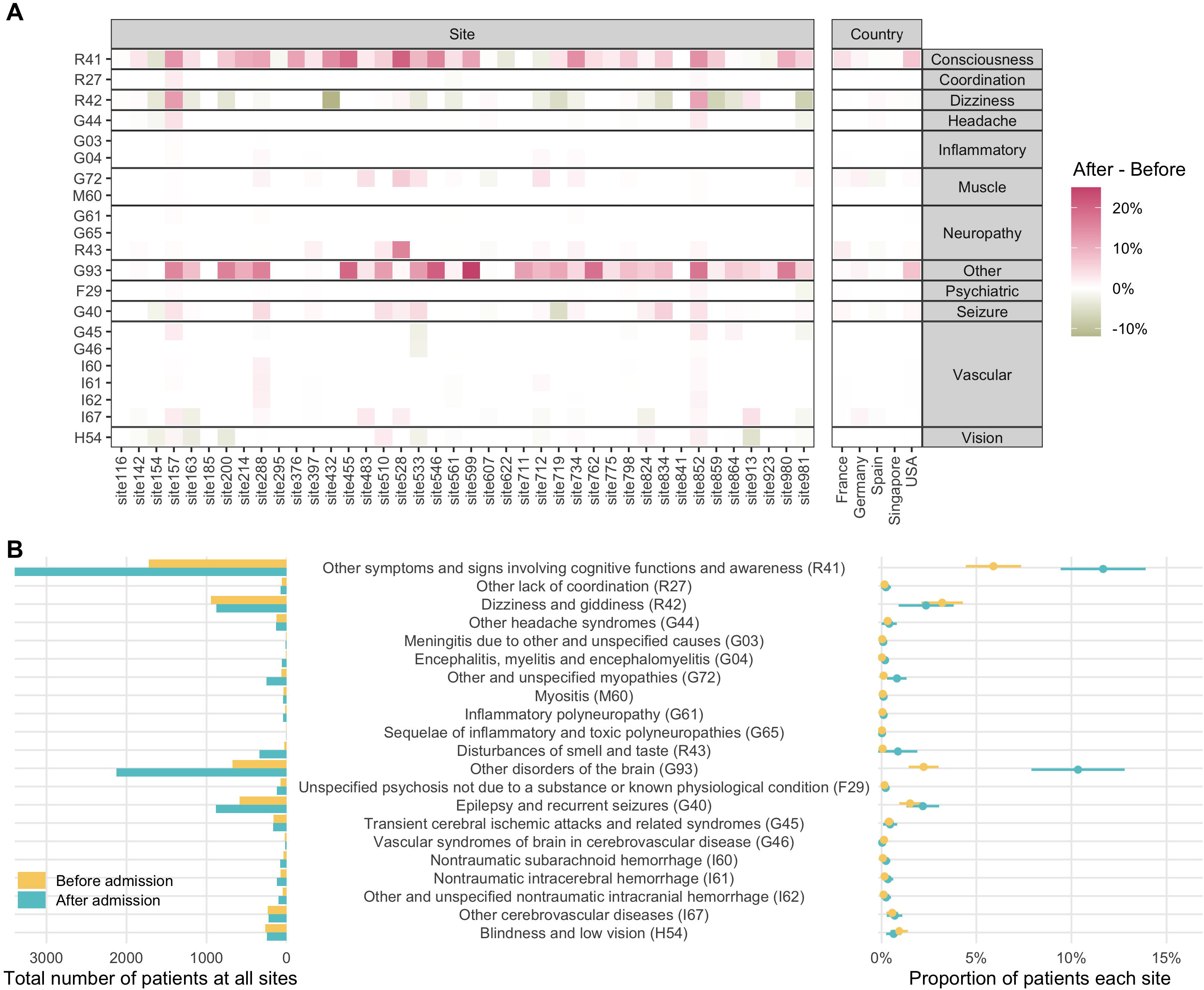
Prevalence of neurological phenotypes among all patients. (**A**) Difference in prevalence of each neurological ICD-10 code by site and country, calculated as after admission - before admission date (**eEq. 2**). Pink color on the heat map indicates increased prevalence, while green color indicates decreased prevalence. Please see **eFig. 1** for the absolute values of prevalence. (**B**) Total counts of patients with a given neurological ICD-10 code (left) and the mean proportion of patients (right) before and after admission date across all sites. The mean proportion estimates are shown as circles and the 95% confidence intervals are shown as bars.

### Prevalence of Neurological Conditions in Severe COVID-19

For each neurological ICD code, we used Fisher’s exact test to examine the enrichment or depletion of the condition among patients who ever experienced severe disease when compared to those who never experienced severe disease (**Fig. 4**). A positive log_2_ value of enrichment (LOE) value denoted a higher proportion of severe cases for a given neurological condition than the never-severe cases, while a negative LOE value indicated the opposite. For instance, a LOE value of 0.283 for ICD code R41 meant that the observed number of severe cases reporting R41 (disorder of consciousness) was 2^0.283^ or ∼1.22 times higher than the expected number of severe cases for R41, which was equivalent to a 22% increase in relative risk (*i.e.,* relative risk difference RRD=22%). We presented in **Table 3** the statistically significant associations of neurological conditions and severe COVID-19 status (*p*_FDR_<0.05). Please refer to the online <u>interactive data table</u> or the <u>results</u> directory of the project online repository for the full table containing the LOE, 95% confidence intervals, and *p* values for all neurological ICD-codes.

**Table 3.** Statistically significant associations of neurological conditions and severe disease status after admission (*p*_FDR_ < 0.05).

| Neurological Condition (ICD-10 Code) | LOE | RRD<sup>a</sup><br>(%) | RRD<sup>a</sup><br>95% CI (%) | $p_{\text{FDR}}$ |
| --- | --- | --- | --- | --- |
| Blindness and low vision (H54) | -0.38 | -23 | (-33, -11) | $2.0 \times 10^{-4}$ |
| Dizziness and giddiness (R42) | -0.47 | -28 | (-33, -22) | $6.3 \times 10^{-19}$ |
| Encephalitis, myelitis and encephalomyelitis (G04) | 0.45 | 37 | (17, 60) | 0.0081 |
| Nontraumatic intracerebral hemorrhage (I61) | 0.45 | 36 | (23, 51) | $3.6 \times 10^{-5}$ |
| Nontraumatic subarachnoid hemorrhage (I60) | 0.35 | 28 | (10, 48) | 0.019 |
| Other and unspecified myopathies (G72) | 0.78 | 72 | (67, 77) | $8.8 \times 10^{-45}$ |
| Other and unspecified nontraumatic intracranial hemorrhage (I62) | 0.43 | 34 | (20, 50) | $3.0 \times 10^{-4}$ |
| Other cerebrovascular diseases (I67) | 0.31 | 24 | (13, 35) | $2.0 \times 10^{-4}$ |
| Other disorders of the brain (G93) | 0.44 | 36 | (32, 40) | $5.6 \times 10^{-73}$ |
| Other headache syndromes (G44) | -0.91 | -47 | (-59, -31) | $9.4 \times 10^{-9}$ |
| Other symptoms and signs involving cognitive functions and awareness (R41) | 0.28 | 22 | (19, 25) | $2.1 \times 10^{-39}$ |
| Transient cerebral ischemic attacks and related syndromes (G45) | -0.85 | -45 | (-56, -31) | $5.0 \times 10^{-10}$ |
| Unspecified psychosis not due to a substance or known physiological condition (F29) | -0.84 | -44 | (-57, -28) | $7.7 \times 10^{-8}$ |

<sup>a</sup>RRD: Relative risk difference = Observed relative risk - 1.

**Figure 4.**
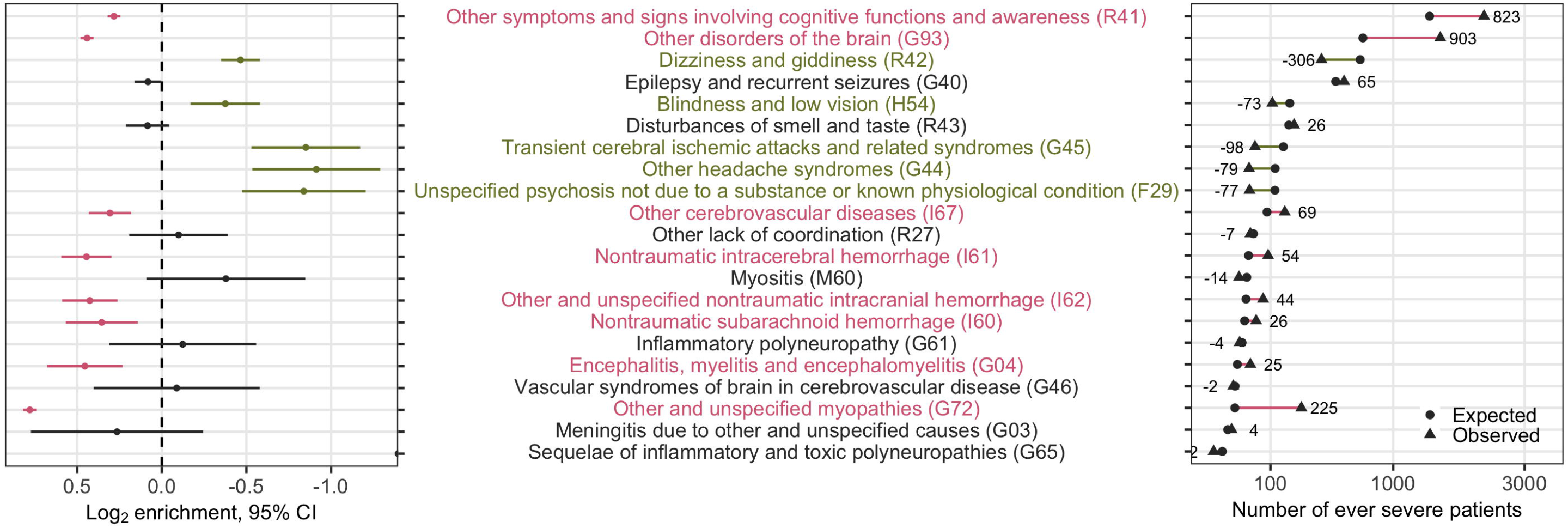
Analysis of enrichment or depletion of neurological conditions after admission in patients with severe disease. For each neurological ICD-10 code, we show the log_2_ enrichment (LOE) and its 95% confidence interval (left), and the absolute difference between the observed (ill) and expected (·) number of patients experiencing severe COVID-19 in square-root scale (right). A **purple** positive LOE value for an ICD-10 code indicates a statistically significantly **higher** proportion of severe cases having a given neurological ICD-10 code when compared to the never-severe cases. Conversely, a **green** negative LOE value indicates a statistically significantly **lower** proportion of severe cases having a given neurological ICD-10 code when compared to the never-severe cases. Neurological ICD-10 codes are ordered by the expected number of severe cases after admission.

In the period after hospital admission date, using the patients who never experienced severe disease as reference, we observed a significantly higher proportion of patients with severe disease who had “other symptoms and signs involving cognitive functions and awareness” (R41: RRD_after_=22%), “other cerebrovascular disease” (ICD-10 I67: RRD_after_=24%), “nontraumatic subarachnoid hemorrhage” (ICD-10 I60: RRD_after_=28%), “other and unspecified nontraumatic intracranial hemorrhage” (ICD-10 I62: RRD_after_=34%), “nontraumatic intracerebral hemorrhage” (ICD-10 I61: RRD_after_=36%), “other disorders of the brain” (ICD-10 G93, including “encephalopathy”: RRD_after_=36%), “encephalitis, myelitis and encephalomyelitis” (ICD-10 G04: RRD_after_=37%), and “other and unspecified myopathies” (ICD-10 G72, including “inflammatory and immune myopathies” and “critical illness myopathies”: RRD_after_=72%). In contrast, we noted a significantly lower proportion of patients with severe disease who had “blindness and low vision” (ICD-10 H54: RRD_after_=-23%), “dizziness and giddiness” (ICD-10 R42: RRD_after_=-28%), “other headache syndromes” (ICD-10 G44: RRD_after_=-47%), “transient cerebral ischemic attacks and related syndromes” (ICD-10 G45: RRD_after_=-45%), and “unspecified psychosis not due to a substance or known physiological condition” (ICD-10 F29: RRD_after_=-44%) during hospitalization (**Table 3, Fig. 4**). Subgroup analysis comparing the US and non-US sites yielded mostly consistent and expected results (**eFig. 2**).

### Exploratory Analysis

As 11 sites in the US and Italy contributed data entirely or partially consisting of ICD-9 codes, we performed separate subgroup analyses using only the ICD-9 data (**eTable 1**), given that one-to-one mapping of some ICD-9 codes to ICD-10 codes was not feasible. To summarize, there was increased prevalence of “disorders of consciousness” and “other neurological conditions” in patients after admission date when compared to before admission date, similar to the ICD-10 data analysis (**eFig. 3, eFig. 4**). The increase in these conditions appear to be driven by the US sites but not the Italian sites. There was no statistically significant difference when examining the change in prevalence of individual neurological ICD-9 codes after admission date. In severity analysis, there were similarities with the ICD-10 data (*e.g.,* “disorders of consciousness”) but also notable differences involving seizure and cerebrovascular events that would require caution in interpretation due to differences in sample size (see **Supplementary Material, eFig. 5, eTable 2**).

## Discussion

In this study, we implemented a research strategy for rapidly aggregating EHR-derived clinical facts across multiple international sites while preserving patient privacy. We report the prevalence of central and peripheral neurological conditions among patients hospitalized with SARS-CoV-2 infection across geographically diverse hospital systems from six countries, using the time period before hospitalization as reference. Importantly, we further compared the prevalence of neurological conditions between patients with and without severe COVID-19 disease.

We found disorders of consciousness and other disorders of the brain as the most prevalent neurological phenotypes (based on ICD codes) among all patients hospitalized with positive SARS-CoV-2, consistent with prior reports.^16,17,21^ Interestingly, our study showed low prevalence of early COVID-19 symptoms such as alterations in smell and taste^12^, likely attributable to the inconsistent documentation of these symptoms in the EHR for the hospitalized patient population, particularly those with severe COVID-19. Further, we did not find statistically significant changes in the prevalence of other previously reported neurological conditions such as dizziness, encephalopathy, headache, seizure, vascular, and vision disorders after admission to the hospital. These discrepancies may be partially due to methodological differences, as our analysis examined the change in prevalence using the data before admission as the reference.

Our major finding indicates that a significantly higher proportion of hospitalized patients with severe COVID-19 had disorders of consciousness, encephalitis and/or myelitis, cerebrovascular events, and myopathy when compared to patients who never had severe disease. While these findings are largely consistent with prior reports^16,20,21^, our contribution is the report of findings from geographically diverse multinational sites. First among these findings, disorders of consciousness, including altered mental status and encephalopathy as well as encephalitis, are consistently reported neurological complications of COVID-19. Sedation for advanced respiratory support and hypoxemia from respiratory failure may partially explain the association of disorders of consciousness with severe COVID-19. Notably, encephalopathy is independently associated with high morbidity and mortality in COVID-19.^21^ Future studies will be important to identify whether the mechanisms underlying encephalitis (with or without myelitis) are direct central nervous system (CNS) invasion by SARS-CoV-2, acute systemic inflammation with secondary CNS involvement, and/or post-infectious immune-mediated effect on the CNS.^28,29^

Second, our finding of cerebrovascular diseases associated with severe COVID-19 include both ischemic strokes and intracranial hemorrhages (nontraumatic intracerebral and nontraumatic subarachnoid), highlighting the difficult balance when managing severe COVID-19 with respect to antiplatelet and anticoagulation therapy. Strokes that occurred in the setting of COVID-19 were associated with high mortality (38% for ischemic stroke and 58% for intracranial hemorrhage)^30^ and morbidity. COVID-19 might increase the risk of ischemic stroke through diverse mechanisms such as activation of innate immune system, cardioembolic events, hypoxia-induced ischemia secondary to severe pulmonary disease, coagulation activation, thrombotic angiopathy, and endothelial damage^31^. Proposed mechanisms underlying intracranial hemorrhage in COVID-19 include coagulation abnormalities, endothelial dysfunction, dysregulation of the renin-angiotensin system, and disruption of cerebral blood flow autoregulation^32,33^. Patients with severe COVID-19 may have additional risk factors such as hypertension or cardiovascular disease that could further drive cerebrovascular diseases.^34^

Finally, while myopathy is common among COVID-19 patients, its mechanism remains unclear. Given that SARS-CoV-2 interacts with the spike domain of ACE2 on host cells^35^ and skeletal muscle tissues express ACE2^36^, muscle may be susceptible to direct SARS-CoV-2 infection. However, myopathy in patients with severe COVID-19 is likely attributable to prolonged or severe critical illness^37^, complications due to multi-organ involvement, or medication-induced myotoxicity (*e.g.,* Hydroxychloroquine, steroids). Our current study design cannot differentiate whether the neurological phenotypes are the direct consequence of SARS-CoV-9 neurotoxicity or due to secondary causes.

Interestingly, some neurological phenotypes are less prevalent in patients with severe COVID-19, including psychosis, dizziness, vision impairment, transient ischemic attack, and headache. The likely explanation is that critically ill patients with or without respiratory failure would either not have objective evaluation or proper documentation and coding for these conditions.

Our study has limitations as the result of trade-offs to standardize data collection from multinational sites while strictly preserving patient privacy and adhering to multinational privacy laws across all contributing sites. First, this study relied on ICD codes that may not capture fully or accurately the disease phenotypes, particularly for conditions better documented in clinical notes. To standardize collection of ICD codes across contributing sites and to mitigate coding discrepancies, we used ICD codes at the categorical level (*e.g.*, the first 3 alphanumeric characters before the decimal point for ICD-10). Thus, further characterization of certain conditions such as “other disorders of the brain” was not feasible at this stage. Second, the proportions of severe cases varied across sites. Adding the number of patients from all sites for each ICD code retained statistical power in the severity analysis, though data from the larger sites likely drove the findings. Third, because we aggregated data across sites, we were unable to consolidate all related ICD codes (*e.g.,* organizing into PheCode^38^) at the individual patient level. The 4CE consortium is addressing these issues to prepare for the next phase of the analyses by developing a framework for conducting patient-level analyses at each site. Fourth, we might not have captured all pre-admission EHR data if patients did not receive all of their care in the same hospital system as the COVID-19 admission. This is a limitation common to all research using EHR data from non-universal health systems. Finally, contributing sites with small patient counts used obfuscation to reduce the risk of re-identifying sites, but the effect of obfuscation is negligible because few sites had neurological conditions of interest in cases fewer than the obfuscation level. Despite the limitations of deploying this rapid, scalable, patient privacy-preserving research strategy, our key findings replicated prior reports from well-characterized but often smaller and single-center cohort studies.

We report the first multinational prevalence study of a broad spectrum of central and peripheral neurological phenotypes in hospitalized patients with PCR-confirmed SARS-CoV-2 infection, particularly those with severe disease, using a common EHR data collection method. Our EHR-based efforts complement registry-based research design. In future studies, we will conduct individual-level analysis using additional EHR data such as complete ICD codes, identify risk factors for neurological phenotypes, and examine long-term neurological sequelae in patients with COVID-19.

## Acknowledgement

We greatly appreciate Margaret Vella in her coordination of the 4CE consortium effort and assistance in the manuscript submission.

## Author Contribution

TL, AGS, JS, GO, PA, SV, DM, and ZX contributed to concept and design of the study. BB, NL, YL, MM, KN, LP, MS, ES, AT, IK, SV, DM, and ZX contributed to data collection. TL, AGS, JS, CH, KN, TC, PA, SV, DM, and ZX contributed to data analysis. TL, AGS, JS, CH, AS, BB, NL, YL, JM, LP, TC, GO, PA, SV, DM, and ZX contributed to drafting and editing the manuscript. All named authors have approved the final manuscript.

