## Supplementary Material for "Multinational Prevalence of Neurological Phenotypes in Patients Hospitalized with COVID-19"

### Prevalence of Neurological Phenotypes in Patients Hospitalized with COVID-19: A Multinational Study of Electronic Health Records

#### Supplemental Materials

##### Supplementary Statistical Methods and Equations

For each neurological ICD code (3 alphanumeric characters), we reported the total count across all sites and all countries (Y) as well as the proportion of patients hospitalized with COVID-19 who had each code at each site (and each country), both before admission ($prop_{before}$) and after admission ($prop_{after}$) (**eQ. 1**).

$$prop_{before}(X,Y) = \frac{number\ of\ hospitalized\ COVID{-}19\ positive\ cases\ diagnosed\ with\ X\ at\ Y before\ admission}{total\ number\ of\ hospitalized\ COVID{-}19\ positive\ cases\ at\ site\ (or\ country)\ Y}\ \textbf{(eEq. 1a)}$$

$$prop_{after}(X,Y) = \frac{number\ of\ hospitalized\ COVID{-}19\ positive\ cases\ diagnosed\ with\ X\ at\ Y after\ admission}{total\ number\ of\ hospitalized\ COVID{-}19\ positive\ cases\ at\ site\ (or\ country)\ Y}\ \textbf{(eEq. 1b)}$$

For each ICD code *X,* we calculated the difference in the proportion of cases with an ICD code *X* before and after COVID-19 hospitalization (**eEq. 2**), and used a paired two-sided t-test to examine whether there was a statistically significant difference when comparing the proportions after admission with proportions before admission across all sites (and countries).

$$\Delta prop(X,Y) = prop_{after}(X,Y) - prop_{before}(X,Y)\ \textbf{(eEq. 2)}$$

We next compared the prevalence of each neurological ICD code (first three characters) and disease category by site, before and after admission date, between patients who ever met the criteria of severe COVID-19 and those who did not. For each ICD code *X*, we computed the expected number of ever-severe patient cases $s_{Ei}$ with:

$$s_{Ei} = \frac{m_X}{\sum_{X\in ICD_{neuro}} m_X} \times \sum_{X\in ICD_{neuro}} s_X\ \textbf{(eEq. 3)}$$

where $m_X$ denotes the observed number of never-severe patient cases with code *X*, and $s_X$ denotes the observed number of ever-severe patient cases with code *X* (**eEq. 3**). Consequently, $\sum_{X\in ICD_{neuro}} s_X$ represents the total number of patients with ever-severe status, and $m_X / \sum_{X\in ICD_{neuro}} m_X$ represents the proportion of patients without severe disease but with neurological code *X*. We then performed an enrichment analysis to examine the difference in proportions of ever-severe disease across neurological ICD codes. Specifically, we calculated the enrichment of each neurological ICD code by dividing the observed number of severe cases by the expected number of severe cases and reported a value of $\log_2$ enrichment (LOE) and its 95% confidence interval (**eEq. 4**).

$$\text{LOE} = log_2 \frac{s_i}{s_{Ei}}\ \textbf{(eEq. 4)}$$

We estimated the 95% confidence interval of the LOE using the Poisson model method. Finally, we computed the *p* values using Fisher's exact test and corrected for multiple hypothesis testing with Benjamini–Hochberg's false discovery rate (FDR) procedure. We considered a result with $p_{FDR}$<0.05 statistically significant.

##### Exploratory Analysis of ICD-9 Data

We analyzed separately the neurological phenotypes among the subset of the patients with ICD-9 codes, as a minority of the 4CE sites in the USA (7 sites) and Italy (4 sites) reported neurological ICD-9 codes (eTable 2). Because 7 of these sites reported both ICD-9 and ICD-10 data and given the aggregate data format, we could not ascertain the exact number of patients with ICD-9 data. We separated this analysis from the main ICD-10 analysis because one-to-one mapping from ICD-9 to ICD-10 codes was not available for all codes.

Similar to the ICD-10 data, there was increased prevalence of "disorders of consciousness and other neurological conditions" in patients after admission date when compared to before admission date (eFig. 2, eFig. 3). These differences appear to be driven by the US sites. However, there was no statistically significant difference when examining the change in prevalence of individual neurological conditions after admission date. The smaller sample size might explain the difference from the ICD-10 data results.

Using the patients who never experienced severe disease as the reference control, in the period after admission date, we found a significantly higher proportion of patients with severe disease to have three broad categorical ICD-9 codes (eFig. 4): (1) "other conditions of the brain" (ICD-9 348, which broadly includes "cerebral cysts", "anoxic brain damage", "benign intracranial hypertension", "encephalopathy, not elsewhere classified", "compression of brain", "cerebral edema": relative risk difference $RRD_{after}$=55%); (2) "general symptoms" (ICD-9 780, which broadly includes "altered consciousness", "hallucination", "syncope and collapse", "convulsions", "dizziness and giddiness", "sleep disturbance", "fever and other physiologic disturbances of temperature regulation", "malaise and fatigue": $RRD_{after}$=33%); (3) "other ill-defined and unknown causes of morbidity and mortality" (ICD-9 799, which broadly includes "asphyxia and hypoxemia", "respiratory arrest", "nervousness", "debility, unspecified", "cachexia", "signs and symptoms involving cognition": $RRD_{after}$=42%). Given the broad definition of these three categorical ICD-9 codes (348, 780, 799), a direct comparison with the ICD-10 data was not feasible and supported the use of ICD-10 data (over ICD-9 data) in the main analysis. Despite the limitations of these ICD-9 codes, the potential presence of "encephalopathy, not elsewhere classified" (a subcode under ICD-9 348), "altered consciousness" (a subcode under ICD-9 780), and "signs and symptoms involving cognition" (a subcode under ICD-9 799) would be consistent with the main findings from the ICD-10 data. Further, "fever and other physiologic disturbances of temperature regulation", "asphyxia and hypoxemia", and "respiratory arrest" would be consistent with severe COVID-19.

Finally, there was a significantly lower proportion of patients with "other and ill-defined cerebrovascular disease" (ICD-9 437: $RDD_{after}$=-42%) among patients with severe disease in the period after admission date. However, we must interpret these findings with great caution given the limitations of the ICD-9 data and the inconsistency with the larger sample size of the ICD-10 data.

**eTable 1. Neurological disease categories, corresponding ICD-9 codes, and their descriptions for the exploratory analysis.**

| Disease Category | ICD-9 | ICD-9 Description |
|---|---|---|
| Muscle | 040 | Other bacterial diseases |
| Psychiatric | 298 | Other nonorganic psychosis |
| Headache | 307 | Special symptoms or syndromes, not elsewhere classified |
| Inflammatory | 320 | Bacterial meningitis |
| Inflammatory | 321 | Meningitis due to other organisms |
| Inflammatory | 322 | Meningitis of unspecified cause |
| Inflammatory | 323 | Encephalitis, myelitis, and encephalomyelitis |
| Other | 330 | Cerebral degenerations usually manifest in childhood |
| Other | 331 | Other cerebral degenerations |
| Headache | 339 | Other headache syndromes |
| Seizure | 345 | Epilepsy and recurrent seizures |
| Other | 348 | Other conditions of brain |
| Neuropathy | 357 | Inflammatory and toxic neuropathy |
| Muscle | 359 | Muscular dystrophies and other myopathies |
| Vision | 369 | Blindness and low vision |
| Vascular | 430 | Subarachnoid hemorrhage |
| Vascular | 431 | Intracerebral hemorrhage |
| Vascular | 432 | Other and unspecified intracranial hemorrhage |
| Vascular | 435 | Transient cerebral ischemia |
| Vascular | 436 | Acute, but ill-defined cerebrovascular disease |
| Vascular | 437 | Other and ill-defined cerebrovascular disease |
| Muscle | 728 | Disorders of muscle, ligament, and fascia |
| Muscle | 729 | Other disorders of soft tissues |
| Other | 780 | General symptoms |
| Neuropathy | 781 | Symptoms involving nervous and musculoskeletal systems |
| Headache | 784 | Symptoms involving head and neck |
| Consciousness | 797 | Senility without mention of psychosis |
| Consciousness | 799 | Other ill-defined and unknown causes of morbidity and mortality |
| Vision/smell/taste | V41 | Problems with special senses and other special functions |

We note that for the four 3-character neurological ICD-9 codes that did not have a one-to-one mapping to corresponding ICD-10 codes, we manually grouped them into different neurological groups:
V41: Problems with special senses and other special functions (Vision/smell/taste)
437: Other and ill-defined cerebrovascular disease (Vascular)
780: General symptoms (Other)
781: Symptoms involving nervous and musculoskeletal systems (Other)

**eTable 2. Statistically significant associations of neurological conditions (ICD-9 codes) after admission and severe disease status ($p_{FDR}$ < 0.05).**

| Neurological Conditions (ICD-9 Code) | LOE | RRD* (%) | RRD* 95% CI (%) | pFDR |
|---|---|---|---|---|
| General symptoms (780) | 0.41 | 33 | (24, 42) | 6.40E-14 |
| Other and ill-defined cerebrovascular disease (437) | -0.76 | -41 | (-60, -14) | 0.022 |
| Other conditions of brain (348) | 0.63 | 55 | (37, 74) | 1.70E-08 |
| Other ill-defined and unknown causes of morbidity and mortality (799) | 0.5 | 42 | (32, 53) | 2.70E-15 |

**RRD: Relative Risk Difference = Observed relative risk - 1.*

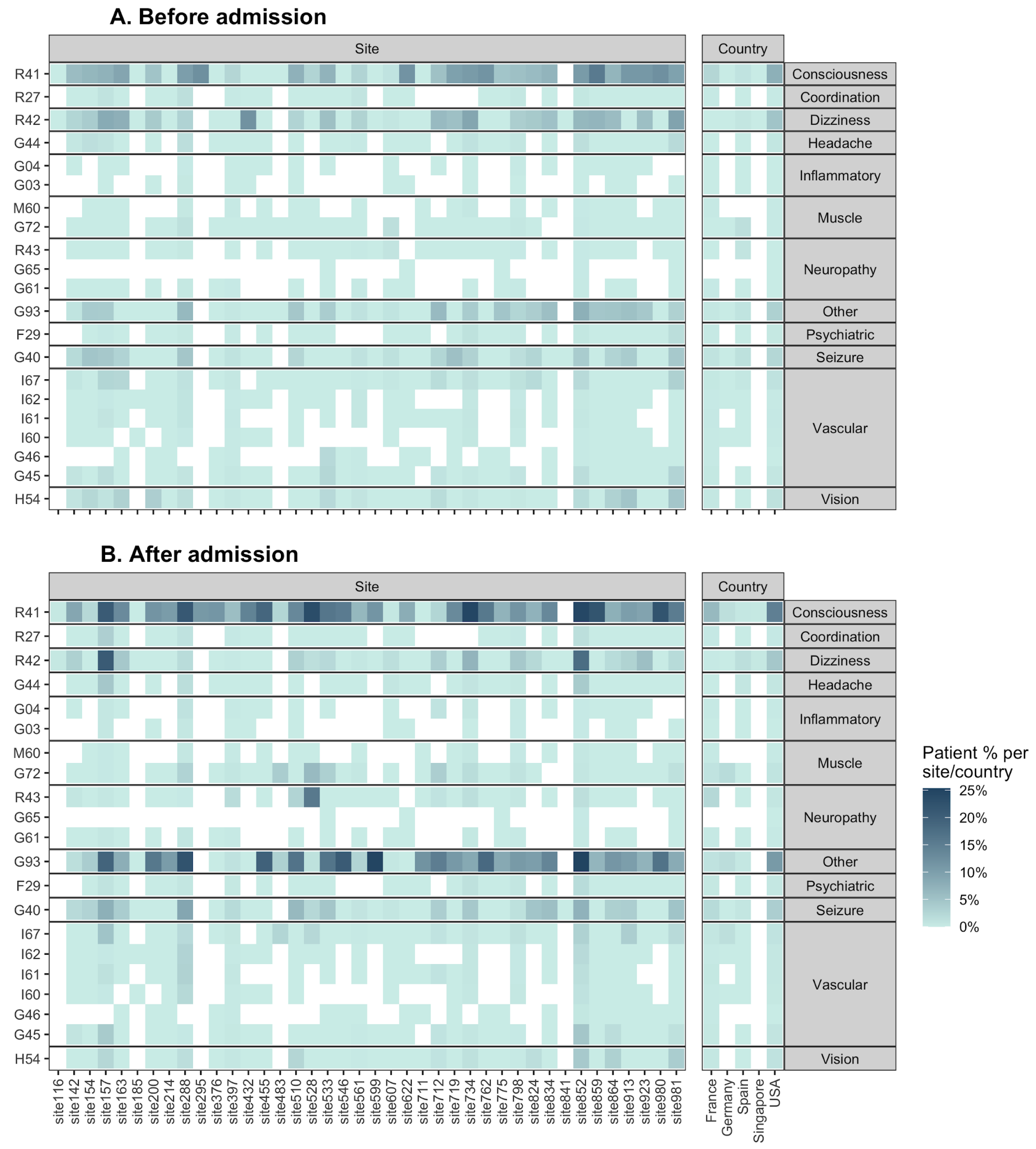

**eFigure 1. Prevalence of each ICD-10 code by site and country before and after admission date, supplementing Fig. 3A.**

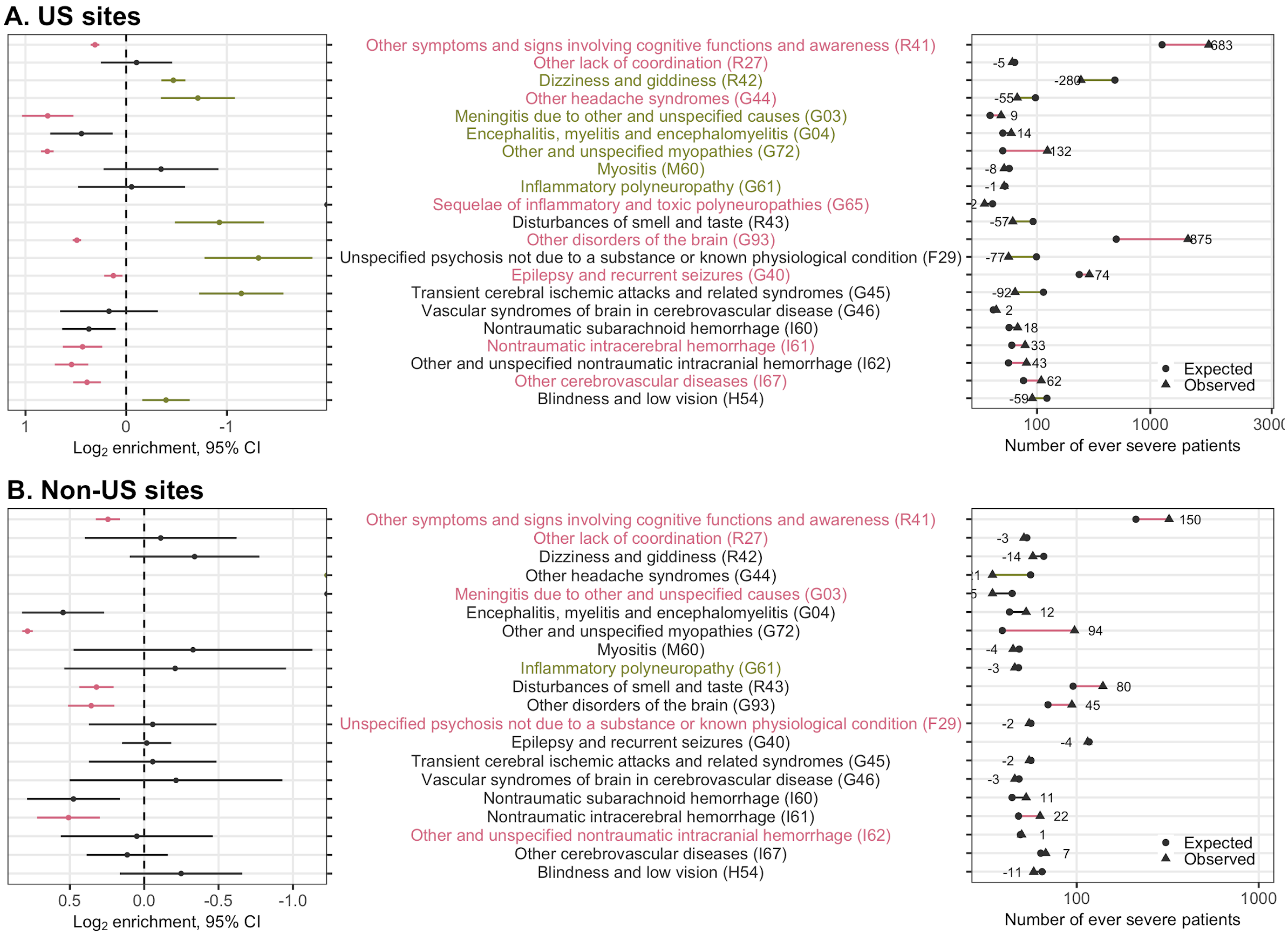

#### eFigure 2. Prevalence of ICD-10 neurological codes among ever-severe cases, grouped by US sites and non-US sites.

$\log_2$ enrichment (LOE) and 95% confidence interval for each ICD-10 code (left) and the absolute difference between the observed (▲) and expected (●) number of severe cases (right) after admission. A **purple** positive LOE value for an ICD-9 code indicates a statistically significantly higher proportion of severe cases with the given ICD-10 code when compared to the never-severe cases. Conversely, a **green** negative LOE value indicates a statistically significantly lower proportion of severe cases with the given ICD-10 code compared to the never-severe cases. Neurological ICD-10 codes are ordered based on the expected number of severe cases after admission date across all sites. The results are generally consistent between the US sites and the non-US sites, except for the following: (1) ICD-10 code R43 (Disturbances of smell and taste) displays opposite directions between the US sites (higher proportion of severe cases with R43) and the non-US sites (lower proportion of severe cases with R43); (2) The US sites have several significant findings that are not significant among the non-US sites, largely due to the smaller number of sites outside the US. Overall, the findings from the subgroup analyses between US and non-US sites are consistent with the findings from the pooled analysis (**Fig. 4**, main text).

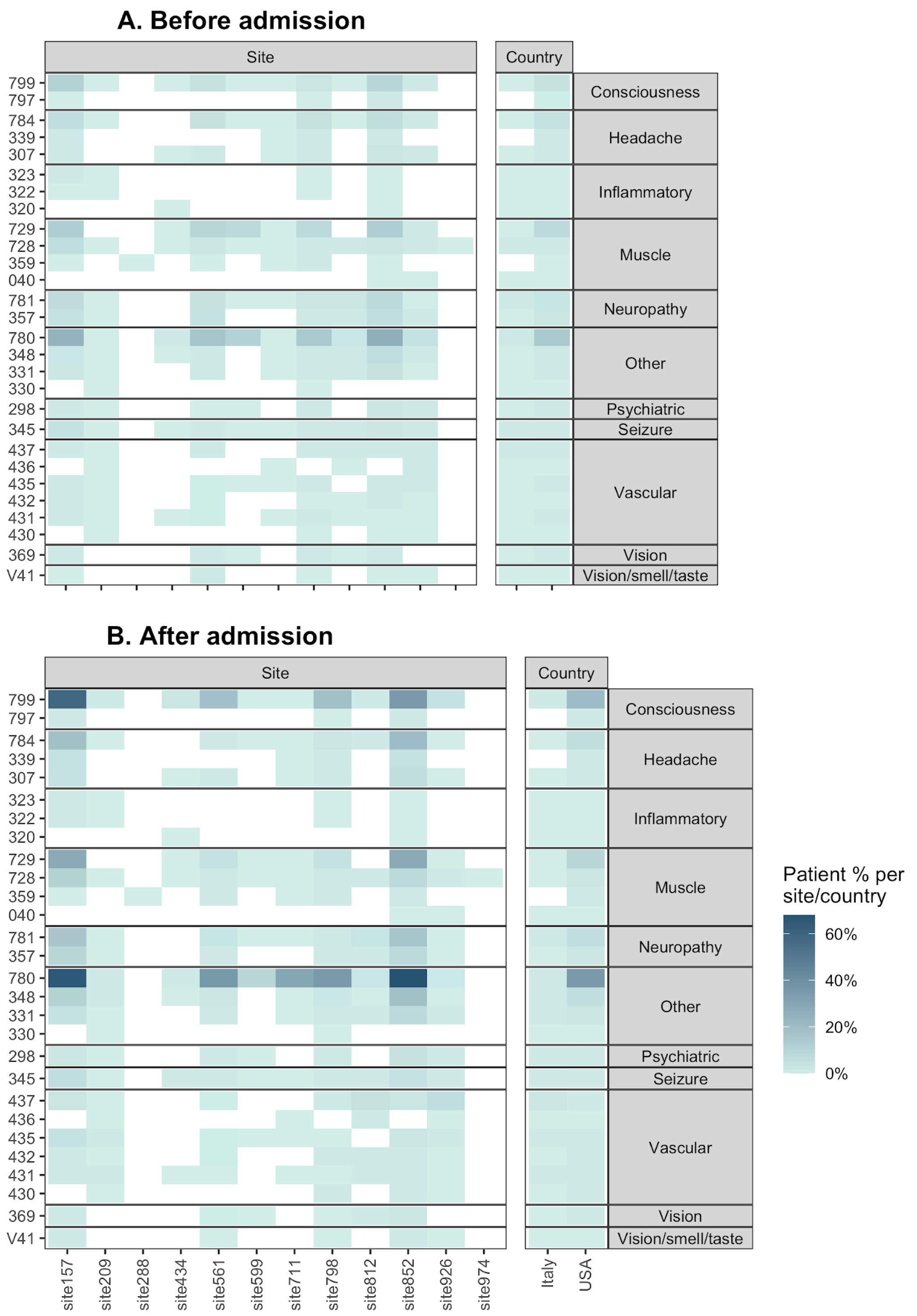

**eFigure 3. Prevalence of each ICD-9 code by site and country before and after admission date.**

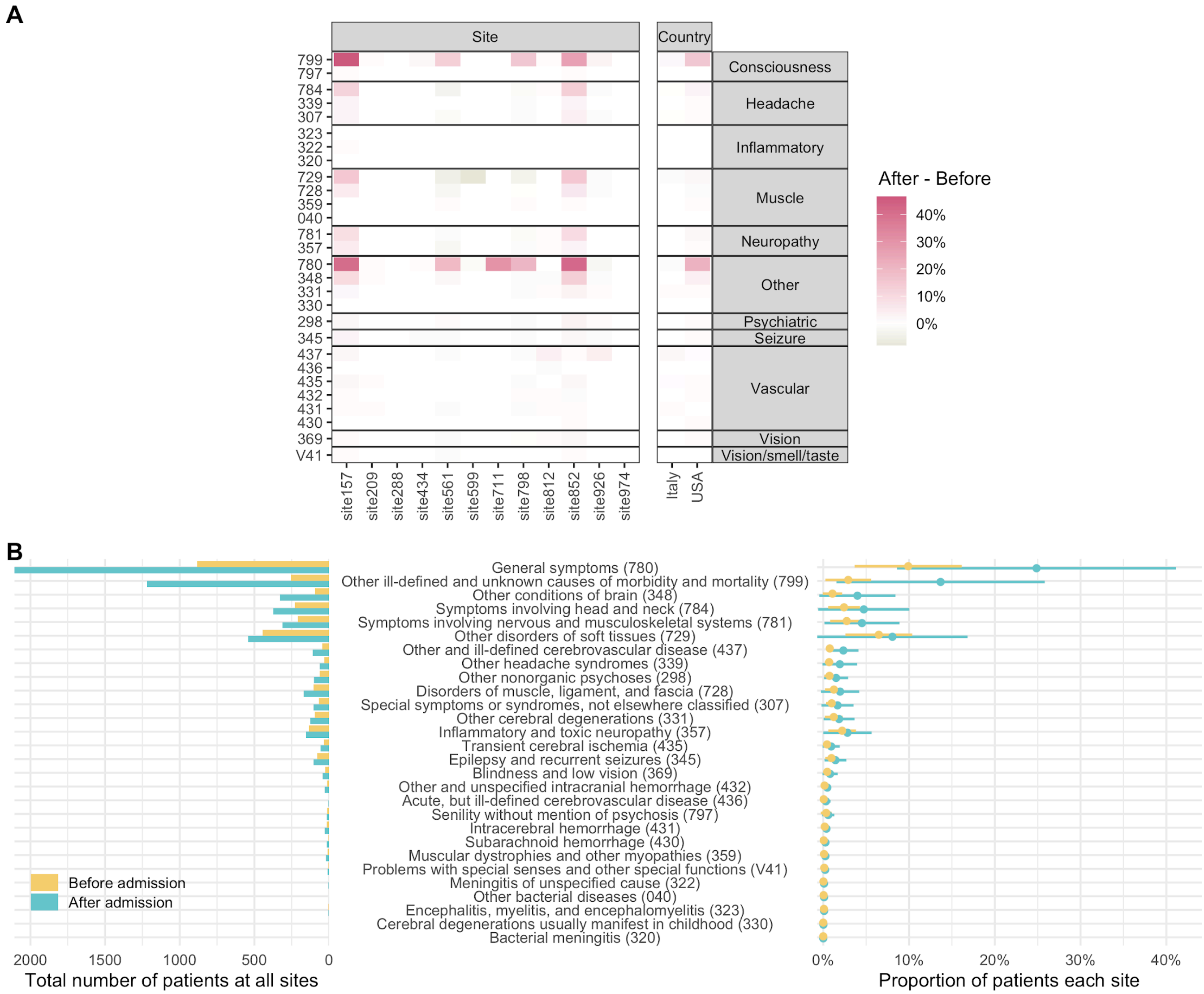

#### eFigure 4. Prevalence of neurological phenotypes among all patients by ICD-9 code.

(A) Difference in prevalence of each neurological ICD-9 code by site and country, calculated as after admission date - before admission date (Eq. 2). The absolute values of prevalence are displayed in **eFig. 3**. (B) Per ICD-9 code, total counts of patients at all sites (left) and average proportion of patients (right) before and after admission date. Mean prevalence estimates across sites are shown as circles and their 95% confidence intervals as bars. ICD-9 codes are ordered based on the mean prevalence difference between before and after admission date.

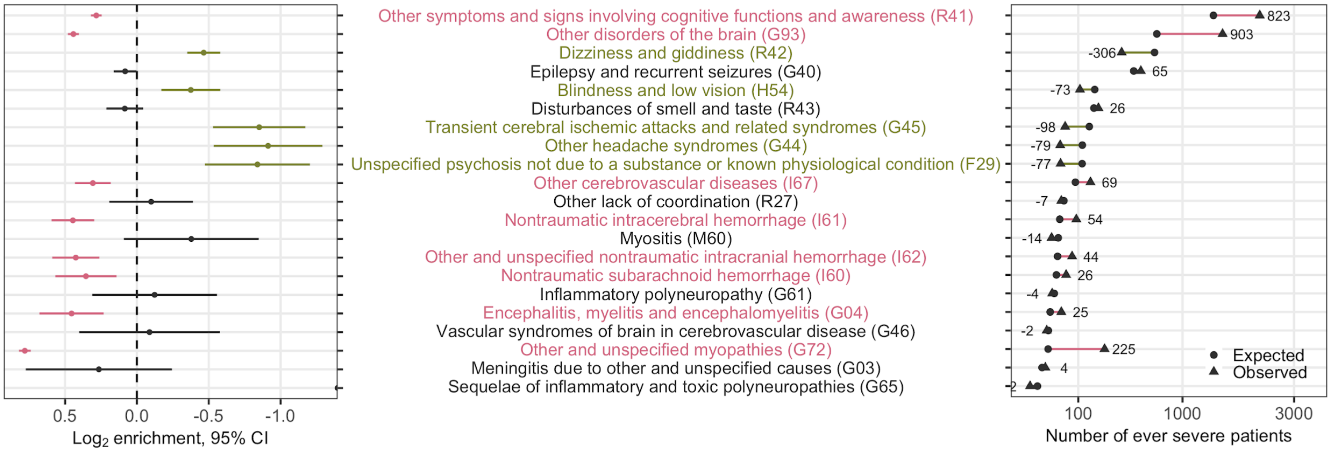

#### eFigure 5. Prevalence of ICD-9 neurological codes among ever-severe cases.

$Log_2$ enrichment (LOE) and 95% confidence interval for each ICD-9 code (left) and the absolute difference between the observed (▲) and expected (●) number of severe cases (right) after admission. A **purple** positive LOE value for an ICD-9 code indicates a statistically significantly higher proportion of severe cases with the given ICD-9 code when compared to the never-severe cases. Conversely, a **green** negative LOE value indicates a statistically significantly lower proportion of severe cases with the given ICD-9 code compared to the never-severe cases. Neurological ICD-9 codes are ordered based on the expected number of severe cases after admission date.
